# Growth mindset and grit as psychological resources in later life: Age, socioeconomic, and health patterning in the English Longitudinal Study of Ageing

**DOI:** 10.64898/2026.02.27.26347198

**Authors:** Paola Zaninotto, Bran Knowles, Jasmine Fledderjohann, Alice Ashcroft, Andrew Steptoe

**Affiliations:** University College London, UK; Lancaster University, UK; Digital Heard Ltd, UK

**Keywords:** growth mindset, grit, psychological resources, older adults

## Abstract

**Objectives:** Growth Mindset and Grit have been proposed as key psychological resources for resilience and adaptation, yet their manifestation and social distribution in later life remain underexplored. This study examines the structure, distribution, and correlates of Growth Mindset and Grit in older adulthood using proxy indicators in the English Longitudinal Study of Ageing (ELSA).

**Methods:** Proxy indicators reflecting learning behaviour, personality traits, affect, and beliefs were used to derive three components of Growth Mindset (education-, personality-, and belief-based) and two components of Grit (affective- and personality-based). Multinomial logistic regression models examined associations with age, socioeconomic position, health, and cognitive functioning.

**Results:** Distinct distributional patterns emerged across components. Education-based Growth Mindset was concentrated in lower categories, whereas personality-, belief-, and affect-based components showed greater variability. Older age was associated with lower Growth Mindset, particularly in education- and belief-based domains, while associations with Grit were more nuanced, including a lower likelihood of low affect-based Grit among older adults. Higher educational attainment, employment, wealth, and better memory performance were associated with more favourable profiles across selected domains. Living alone and limiting longstanding illness were consistently associated with less favourable profiles.

**Conclusions:** Growth Mindset and Grit appear to function as multidimensional and socially patterned psychological resources in later life. Belief-, personality-, and affect-based components capture meaningful variation even when formal learning declines, underscoring the importance of distinguishing opportunity-constrained indicators from dispositional domains in ageing research.

## Introduction

Psychological resources that support learning, motivation, and persistence are increasingly recognised as important for understanding how individuals adapt to challenges across the life course. In later life, when opportunities, constraints, and vulnerabilities become more heterogeneous, such resources may be particularly relevant for explaining variation in engagement, resilience, and wellbeing among older adults. Two constructs that have attracted growing attention in this context are growth mindset and grit, which reflect orientations toward change, effort, and long-term goal pursuit. Despite their theoretical relevance to ageing, however, evidence on how these constructs are distributed and patterned across the older population remains limited.

A growth mindset refers to the belief that abilities and personal characteristics are malleable and can be developed through effort and learning (Dweck, 2006). While most research has focused on educational settings, growth-oriented beliefs may be particularly salient in later life, where age-related stereotypes and expectations of decline could shape attitudes toward learning, adaptation, and self-improvement (Dweck & Yeager, 2019; Heslin et al., 2021). Existing studies in older adults - largely cross-sectional and based on modest or regionally specific samples - suggest that growth mindset is associated with health behaviours, cognitive performance, and indicators of successful ageing. Intervention studies further indicate that growth-oriented beliefs remain malleable in later life. For example, older adults participating in multi-skill learning programmes have shown cognitive gains associated with individual differences in growth mindset (Sheffler et al., 2023), and structured interventions designed to foster growth-oriented attitudes have demonstrated improvements in cognitive functioning (Leanos et al., 2023). However, such studies typically involve selected volunteers and are not designed to characterise population-level distribution.

Alongside growth mindset, the construct of grit has been proposed as a related psychological resource reflecting perseverance of effort and consistency of interests over time (Duckworth, 2016). Whereas growth mindset concerns beliefs about the possibility of change, grit captures sustained effort toward long-term goals. In later life, grit may be particularly relevant for understanding how older adults maintain engagement and adapt to age-related challenges, although its role in ageing remains under-examined.

Research on grit in older adulthood has largely relied on cross-sectional survey designs, often using self-report measures in relatively small or regionally specific samples. For example, a study examined grit and successful ageing in a community-based sample of 315 older adults in Korea (Kim & Lee, 2015), reporting positive associations between perseverance and successful ageing indicators. A US study using a sample of 185 older adults (Rhodes & Giovannetti, 2022), found that grit was associated with multiple domains of physical, emotional, and social functioning. Fleck et al. (2024) similarly reported associations between grit and health outcomes in older adults. While these studies consistently suggest that grit is relevant to ageing-related outcomes, their designs do not allow for an assessment of how grit is socially patterned across the full spectrum of later-life experiences at the population level.

Taken together, the existing literature provides important but partial insights. Evidence remains limited from large, nationally representative ageing cohorts that can situate growth mindset and grit within broader social, health, and cognitive contexts. Large population-based studies offer an opportunity to examine these psychological dispositions not only as individual traits but as outcomes shaped by socioeconomic position, health status, and cognitive functioning.

The present study uses data from the English Longitudinal Study of Ageing (ELSA), (Steptoe et al., 2013), a nationally representative cohort of older adults in England, to operationalise proxy indicators of growth mindset and grit and to examine how these constructs are distributed across later life. By providing a population-level perspective, this study extends prior research based largely on smaller or more selective samples and contributes to understanding the social and health patterning of growth-oriented psychological resources in older adulthood.

## Methods

### Data

The data used in this study come from the English Longitudinal Study of Ageing (ELSA) an interdisciplinary ongoing cohort study of older adults aged 50 years and older living in private households in England (Steptoe et al., 2013). The study began in 2002 with a sample of approximately 12,000 individuals (response rate 67%). Data are collected biennially using face-to-face personal interviews, self-completion questionnaires and health visits (every four years). The sample is periodically refreshed with new participants to ensure the study remains representative of those aged 50 and over. Details of the survey’s sampling frame, methodology, and questionnaires can be found at www.elsa-project.ac.uk. For the purpose of this study we used data from Wave 5 (2010-11) when relevant information where first collected. A total 9,090 participants who completed the survey in Wave 5.

ELSA was approved by the London Multicentre Research Ethics Committee (MREC/01/2/91). Informed consent was obtained from all participants. All data are available through the UK Data Service (SN 5050).

## Measures

Growth mindset and grit were operationalised using theory-informed proxy indicators derived from personality, belief, behavioural, and affective measures available in Wave 5 of ELSA. All items were reverse-coded where necessary and standardised (z-scores) prior to index construction. Continuous composite scores were created by averaging standardised items and categorised into low (< −0.5 SD), moderate (−0.5 to 0.5 SD), and high (> 0.5 SD) groups. (see Supplementary materials for full description and Supplementary Table 1 for full item wording).

### Growth Mindset Personality

Trait-based growth mindset was derived from five items (Curious, Imaginative, Intelligent, Creative, Broad-minded) from the Midlife Development Inventory (MIDI)(Lachman & Weaver, 1997), reflecting openness to change and intellectual engagement.

### Growth Mindset Beliefs

Growth-oriented beliefs were operationalised using three CASP-19 items (Hyde et al., 2003) capturing perceived opportunity, future orientation, and willingness to engage in new experiences.

### Growth Mindset Adult Learning

Behavioural engagement in growth-oriented learning was measured using participation in formal education or training across Waves 4-5, categorised according to degree of engagement over the preceding four years.

### Grit Personality

Personality-based grit was derived from five MIDI items (Hardworking, Responsible, Calm, Organised, Active, Thorough) reflecting perseverance, focus, and diligence.

### Grit Affect

Affect-based grit was constructed from five positive affect items (Determined, Hopeful, Attentive, Interested, Inspired) from the (PANAS-X; Watson & Clark, 1994), reflecting motivational engagement and sustained effort.

## Individual characteristics

Several variables were included to describe the sample characteristics for each growth mindset and grit measure. Socio-demographic characteristics comprised age, sex (male or female), living arrangement (alone vs not alone), ethnicity (White vs Non-White), age finished full-time education (14 or under, 15 to 18, 19 or over), employment status (in work, retired, other) and wealth tertiles (measured as the sum of all physical wealth minus debts). Health was assessed using a question on self-reported limiting longstanding illness asking whether the participant “has any long-standing illness, disability, or infirmity that has troubled [them] over a period of time or is likely to affect [them] over a period of time.” Participants responding affirmatively were classified as having a limiting longstanding illness. We also included a cognitive function score derived from a memory assessment. Memory was evaluated using a word-list learning test, in which participants were presented orally with a list of ten words to recall immediately and after a short delay (Zaninotto et al., 2018). The overall memory score ranged from 0 to 20.

## Statistical analysis

The analytical sample consisted of individuals who completed the main interview and the self-completion questionnaire in Wave 5 (8,075 out of the 9,090 total participants in that wave 89%). Complete-case analysis was carried and we therefore selected people with valid data on all variables of interest (total of 7,333 out of 8,075, 91%).

Descriptive statistics and regression models were weighted for non-response using inverse probability sampling weights. All analyses were conducted in Stata v19.

## Results

### Descriptive statistics

Supplementary Table 2 shows the distribution of growth mindset and grit components. Growth mindset based on adult education was heavily skewed towards the low category, whereas personality and belief components were more evenly distributed. Grit components showed broadly balanced distributions across low, moderate, and high categories.

Supplementary Table 3 indicates age gradients across most components, particularly for adult education-based growth mindset, where participants in the low group were substantially older than those in the high group. Age differences were more modest for personality and affect-based components.

### Adult education component of growth mindset

Table 1 presents the relative risk ratios (RRRs) and 95% confidence intervals (CIs) from a multinomial logistic regression model examining simultaneously socio-demographic and health-related predictors of the growth mindset-oriented adult education component.

**Table 1.**
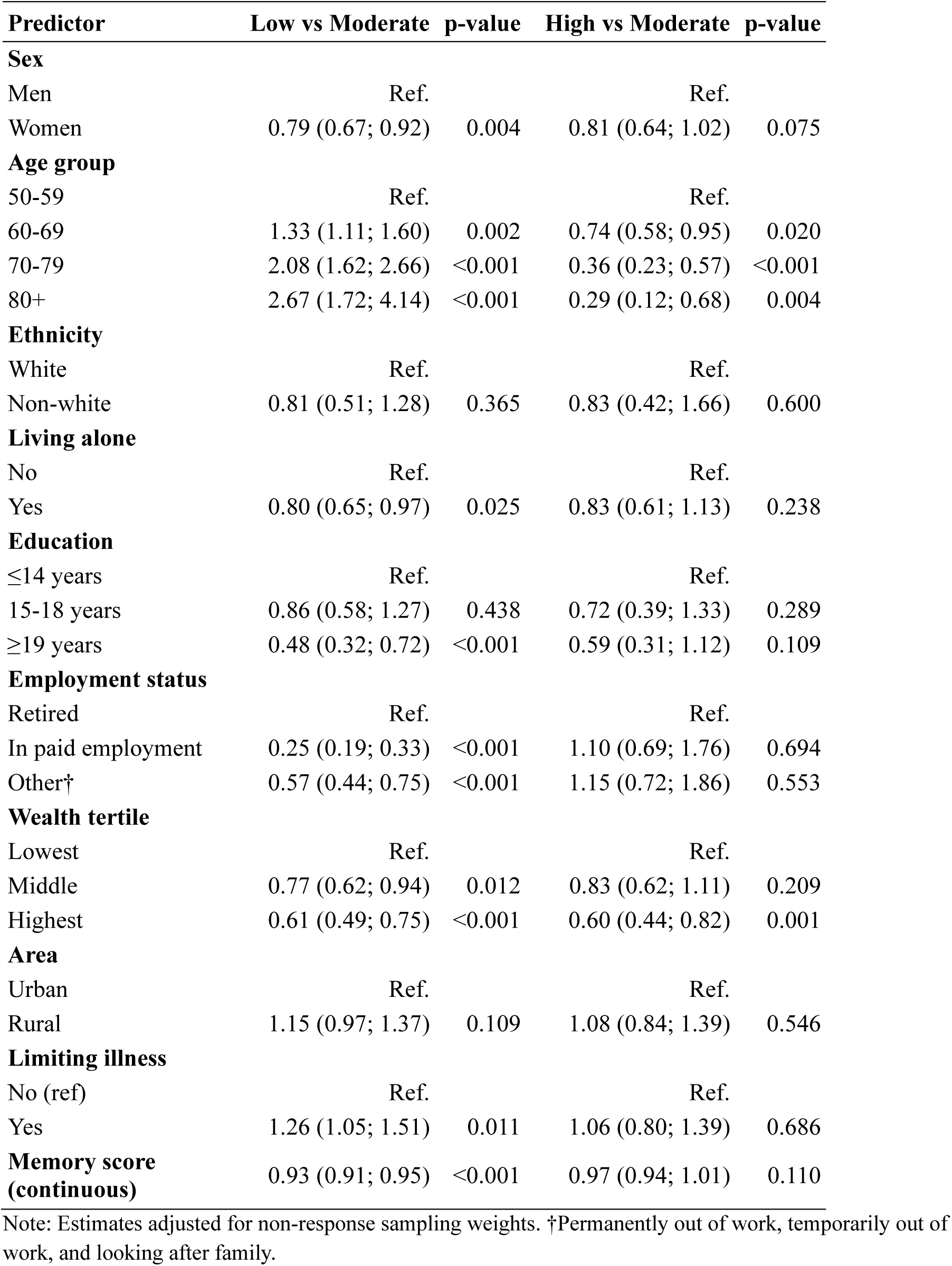
Predictors of the growth mindset-oriented adult education component, England 2008-2012.

| **Predictor** | **Low vs Moderate** | **p-value** | **High vs Moderate** | **p-value** |
| --- | --- | --- | --- | --- |
| **Sex** |  |  |  |  |
| Men | Ref. |  | Ref. |  |
| Women | 0.79 (0.67; 0.92) | 0.004 | 0.81 (0.64; 1.02) | 0.075 |
| **Age group** |  |  |  |  |
| 50-59 | Ref. |  | Ref. |  |
| 60-69 | 1.33 (1.11; 1.60) | 0.002 | 0.74 (0.58; 0.95) | 0.020 |
| 70-79 | 2.08 (1.62; 2.66) | <0.001 | 0.36 (0.23; 0.57) | <0.001 |
| 80+ | 2.67 (1.72; 4.14) | <0.001 | 0.29 (0.12; 0.68) | 0.004 |
| **Ethnicity** |  |  |  |  |
| White | Ref. |  | Ref. |  |
| Non-white | 0.81 (0.51; 1.28) | 0.365 | 0.83 (0.42; 1.66) | 0.600 |
| **Living alone** |  |  |  |  |
| No | Ref. |  | Ref. |  |
| Yes | 0.80 (0.65; 0.97) | 0.025 | 0.83 (0.61; 1.13) | 0.238 |
| **Education** |  |  |  |  |
| ≤14 years | Ref. |  | Ref. |  |
| 15-18 years | 0.86 (0.58; 1.27) | 0.438 | 0.72 (0.39; 1.33) | 0.289 |
| ≥19 years | 0.48 (0.32; 0.72) | <0.001 | 0.59 (0.31; 1.12) | 0.109 |
| **Employment status** |  |  |  |  |
| Retired | Ref. |  | Ref. |  |
| In paid employment | 0.25 (0.19; 0.33) | <0.001 | 1.10 (0.69; 1.76) | 0.694 |
| Other† | 0.57 (0.44; 0.75) | <0.001 | 1.15 (0.72; 1.86) | 0.553 |
| **Wealth tertile** |  |  |  |  |
| Lowest | Ref. |  | Ref. |  |
| Middle | 0.77 (0.62; 0.94) | 0.012 | 0.83 (0.62; 1.11) | 0.209 |
| Highest | 0.61 (0.49; 0.75) | <0.001 | 0.60 (0.44; 0.82) | 0.001 |
| **Area** |  |  |  |  |
| Urban | Ref. |  | Ref. |  |
| Rural | 1.15 (0.97; 1.37) | 0.109 | 1.08 (0.84; 1.39) | 0.546 |
| **Limiting illness** |  |  |  |  |
| No (ref) | Ref. |  | Ref. |  |
| Yes | 1.26 (1.05; 1.51) | 0.011 | 1.06 (0.80; 1.39) | 0.686 |
| **Memory score (continuous)** | 0.93 (0.91; 0.95) | <0.001 | 0.97 (0.94; 1.01) | 0.110 |

Note: Estimates adjusted for non-response sampling weights. †Permanently out of work, temporarily out of work, and looking after family.

#### Low growth mindset adult education component versus moderate

After controlling for all covariates, women were less likely than men to report a low personality-based growth mindset (RRR = 0.79, 95% CI: 0.67; 0.92, p = 0.004). Age showed a strong graded association, with progressively higher relative risks of low growth mindset at older ages compared with those aged 50-59 (RRR = 1.33 for ages 60-69; 2.08 for 70-79; and 2.67 for 80+, all p < 0.01). Individuals living alone were less likely to report a low growth mindset than those not living alone (RRR = 0.80, 95% CI: 0.65; 0.97, p = 0.025). Higher educational attainment was protective: participants who completed education at age 19 years or older were substantially less likely to report a low growth mindset compared with those who left school before age 14 (RRR = 0.48, 95% CI: 0.32; 0.72, p < 0.001). Employment status was a strong predictor: individuals in paid employment had markedly lower relative risks of low growth mindset compared with retirees (RRR = 0.25, 95% CI: 0.19; 0.33, p < 0.001), with similarly reduced risks among those in other non-retired employment categories (RRR = 0.57, 95% CI: 0.44; 0.75, p < 0.001). Higher household wealth was also associated with a lower likelihood of low growth mindset.

There was not enough evidence that ethnicity or urbanicity were associated with low growth mindset after accounting for other predictors. There was also not enough evidence that memory score was independently associated with low growth mindset once other characteristics were considered.

#### High growth mindset adult education versus moderate

In contrast, after accounting for other predictors, sex, living arrangement, educational attainment, employment status, limiting longstanding illness, ethnicity, and urbanicity did not distinguish individuals with a high growth mindset from those with a moderate growth mindset. Older age and higher wealth, however, were indicative of differences: compared with those aged 50-59, participants in older age groups were progressively less likely to report a high growth mindset, and those in the highest wealth group were also less likely to report a high (rather than moderate) growth mindset.

### Personality component of growth mindset

Table 2 presents the predictors of the personality component of the growth mindset, explored simultaneously in the model.

**Table 2.**
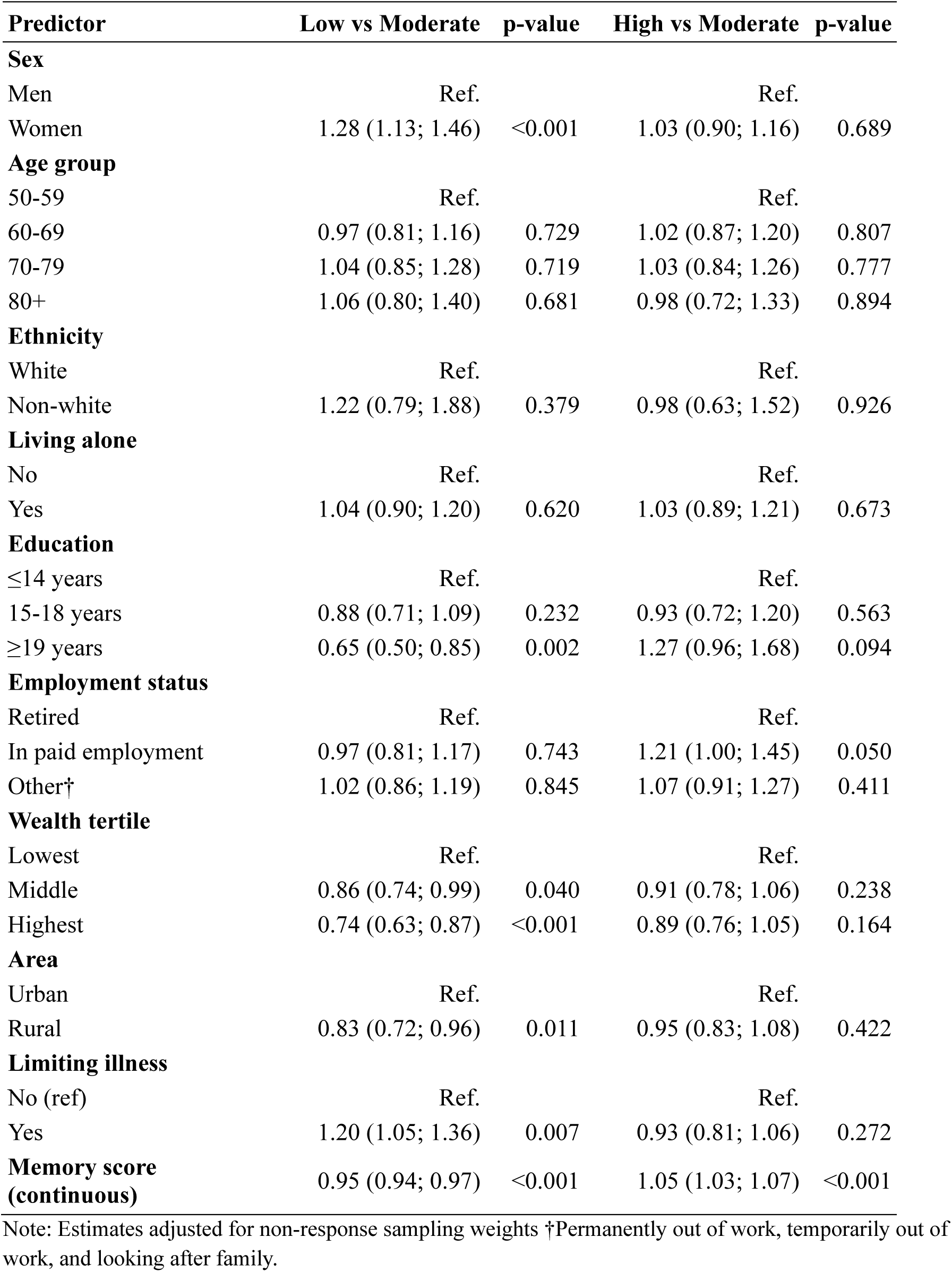
Predictors of the growth mindset-oriented personality component, England 2008-2012.

#### Low growth mindset personality versus moderate

After accounting for all other characteristics, females were significantly more likely than males to report a low personality-based growth mindset (RRR = 1.28, 95% CI: 1.13; 1.46, p < 0.001). Age was not significantly associated with the likelihood of reporting a low growth mindset compared to a moderate one. Ethnicity and living alone were not related to a higher relative risk of low growth mindset compared to medium. Educational attainment showed a clear protective effect: those who completed education at age 19 years or older were substantially less likely to report a low growth mindset compared with those who left school before age 14 (RRR = 0.65, 95% CI: 0.50; 0.85, p < 0.001). Employment status was not significantly associated with growth mindset. Higher household wealth and living in rural areas were associated with a lower likelihood of low growth mindset. Participants with a limiting longstanding illness were more likely to report a low growth mindset (RRR = 1.20, 95% CI: 1.05; 1.39, p = 0.007), whereas higher memory scores were strongly associated with a lower likelihood of low growth mindset (RRR = 0.95 per unit increase, 95% CI: 0.94; 0.97, p < 0.001).

#### High personality growth mindset personality versus moderate

In contrast, none of the characteristics revealed a significant likelihood of being in the high growth mindset group compared to moderate, with the exception of memory.

Higher memory scores were positively associated with a high growth mindset (RRR = 1.05 per unit increase, 95% CI: 1.03; 1.07, p < 0.001).

### Beliefs component of growth mindset

Table 3 presents the predictors of the growth mindset oriented beliefs component, explored simultaneously in the model.

**Table 3.** Predictors of the growth mindset-oriented belief component, England 2008-2012.

| **Predictor** | **Low vs Moderate** | **p-value** | **High vs Moderate** | **p-value** |
| --- | --- | --- | --- | --- |
| **Sex** |  |  |  |  |
| Men | Ref. |  | Ref. |  |
| Women | 0.89 (0.78; 1.02) | 0.086 | 1.05 (0.93; 1.18) | 0.442 |
| **Age group** |  |  |  |  |
| 50-59 | Ref. |  | Ref. |  |
| 60-69 | 0.83 (0.68; 1.00) | 0.050 | 1.08 (0.92; 1.26) | 0.361 |
| 70-79 | 0.89 (0.72; 1.10) | 0.286 | 0.86 (0.71; 1.04) | 0.121 |
| 80+ | 1.28 (0.95; 1.71) | 0.104 | 0.74 (0.55; 1.00) | 0.046 |
| **Ethnicity** |  |  |  |  |
| White | Ref. |  | Ref. |  |
| Non-white | 0.84 (0.51; 1.38) | 0.494 | 1.01 (0.66; 1.54) | 0.961 |
| **Living alone** |  |  |  |  |
| No | Ref. |  | Ref. |  |
| Yes | 1.43 (1.23; 1.66) | <0.001 | 0.89 (0.76; 1.03) | 0.122 |
| **Education** |  |  |  |  |
| ≤14 years | Ref. |  | Ref. |  |
| 15-18 years | 1.02 (0.81; 1.29) | 0.854 | 1.03 (0.81; 1.31) | 0.807 |
| ≥19 years | 0.86 (0.65; 1.15) | 0.311 | 1.16 (0.88; 1.52) | 0.288 |
| **Employment status** |  |  |  |  |
| Retired | Ref. |  | Ref. |  |
| In paid employment | 0.71 (0.58; 0.86) | 0.001 | 0.95 (0.80; 1.14) | 0.596 |
| Other† | 0.98 (0.83; 1.16) | 0.827 | 0.97 (0.82; 1.14) | 0.716 |
| **Wealth tertile** |  |  |  |  |
| Lowest | Ref. |  | Ref. |  |
| Middle | 0.74 (0.64; 0.87) | <0.001 | 1.09 (0.93; 1.26) | 0.278 |
| Highest | 0.61 (0.51; 0.72) | <0.001 | 1.47 (1.26; 1.71) | <0.001 |
| **Area** |  |  |  |  |
| Urban | Ref. |  | Ref. |  |
| Rural | 0.96 (0.83; 1.12) | 0.623 | 1.07 (0.94; 1.21) | 0.317 |
| **Limiting illness** |  |  |  |  |
| No (ref) | Ref. |  | Ref. |  |
| Yes | 2.22 (1.94; 2.54) | <0.001 | 0.61 (0.53; 0.70) | <0.001 |
| **Memory score (continuous)** | 0.98 (0.96; 1.00) | 0.054 | 1.05 (1.03; 1.07) | <0.001 |

Note: Estimates adjusted for non-response sampling weights. †Permanently out of work, temporarily out of work, and looking after family.

#### Low growth mindset beliefs versus moderate

After controlling for all other variables, there was not enough evidence that sex was associated with low growth mindset beliefs. Compared with those aged 50-59, participants aged 60-69 were less likely to report low beliefs (RRR = 0.83, 95% CI: 0.68; 1.00, p = 0.050), while those aged 70-79 and 80+ did not differ significantly. Individuals living alone were substantially more likely to report low growth mindset beliefs compared with those not living alone (RRR = 1.43, 95% CI: 1.23; 1.66, p < 0.001). There was not enough evidence that educational attainment was associated with low beliefs after adjustment. Being in paid employment was associated with a lower likelihood of low growth mindset beliefs compared with being retired (RRR = 0.71, 95% CI: 0.58; 0.86, p = 0.001), while other non-retired groups did not differ significantly. Household wealth showed a clear gradient: compared with the lowest tertile, those in the middle (RRR = 0.74, 95% CI: 0.64; 0.87, p < 0.001) and highest tertiles (RRR = 0.61, 95% CI: 0.51; 0.72, p < 0.001) were less likely to report low growth mindset beliefs. There was not enough evidence that ethnicity or urban-rural residence were associated with low beliefs. Participants with a limiting longstanding illness had a markedly higher likelihood of low beliefs (RRR = 2.22, 95% CI: 1.94; 2.54, p < 0.001). Higher memory scores were associated with a lower likelihood of low growth mindset beliefs (RRR = 0.98 per unit increase, 95% CI: 0.96; 1.00, p = 0.054), although this association was borderline in strength.

#### High growth mindset beliefs versus moderate

After controlling for all other variables, most characteristics were not associated with the likelihood of being in the high growth mindset beliefs group compared with the moderate group. The main exceptions were wealth, limiting longstanding illness, and memory. Household wealth was strongly associated: compared with the lowest tertile, those in the highest wealth tertile were more likely to report high growth mindset beliefs (RRR = 1.47, 95% CI: 1.26; 1.71, p < 0.001), while the middle tertile did not differ significantly. Having a limiting longstanding illness was associated with a lower likelihood of high growth mindset beliefs (RRR = 0.61, 95% CI: 0.53; 0.70, p < 0.001). Higher memory scores were positively associated with high growth mindset beliefs (RRR = 1.05 per unit increase, 95% CI: 1.03; 1.07, p < 0.001).

### Affect Grit

Table 4 presents the relative risk ratios (RRRs) and 95% confidence intervals (CIs) from a multinomial logistic regression model examining simultaneously socio-demographic and health-related predictors of grit-oriented affect. The reference category for the outcome was moderate grit.

**Table 4.**
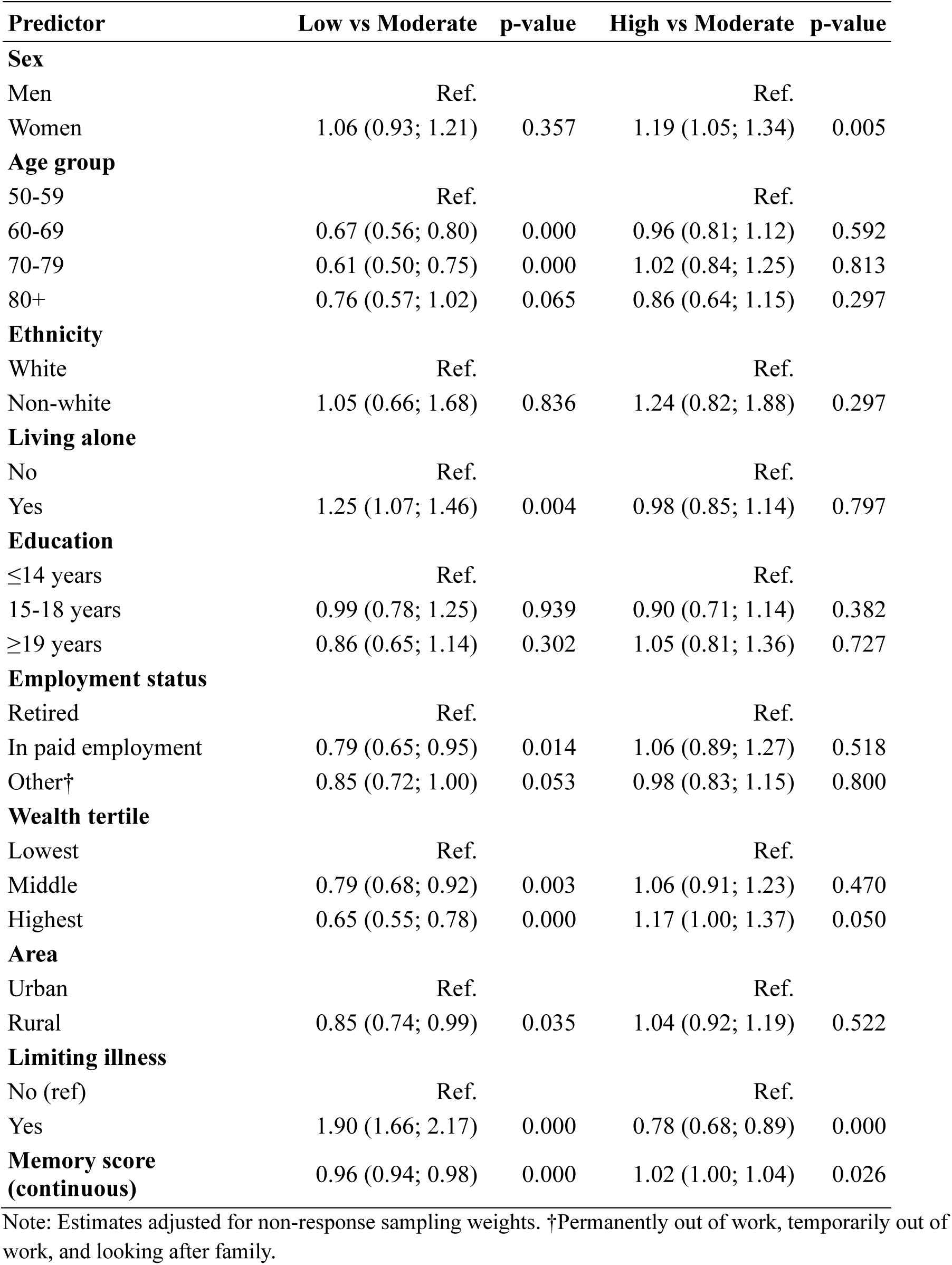
Predictors of the grit-oriented affect component, England 2008-2012.

#### Low affect grit versus moderate

After accounting for all other characteristics, participants aged 60-69, 70-79, and 80+ were significantly less likely to have low compared with moderate grit (RRR = 0.67, 0.61, and 0.76, respectively; p < 0.01 for 60-69 and 70-79, p = 0.065 for 80+). Living alone was associated with a higher likelihood of low grit (RRR = 1.25, 95% CI 1.07; 1.46), as was reporting a limiting illness (RRR = 1.90, 95% CI 1.66; 2.17). Gender and ethnicity were not significantly associated with low grit. Being in paid employment was linked to lower odds of low grit (RRR = 0.79, 95% CI 0.65; 0.95). Higher education and wealth were associated with lower relative risk of low grit: middle wealth tertile (RRR = 0.79, 95% CI 0.68; 0.92), highest wealth tertile (RRR = 0.65, 95% CI 0.55-0.78). Living in a rural area was associated with slightly lower relative risk of low grit (RRR = 0.85, 95% CI 0.74; 0.99). Better memory performance was associated with reduced relative risk of low grit (RRR = 0.96, 95% CI 0.94; 0.98).

#### High affect grit versus moderate

When comparing high versus moderate grit, few characteristics seemed to matter. For example, women were more likely than men to report high grit (RRR = 1.19, 95% CI 1.05; 1.34). Age, ethnicity and living alone were not significantly associated with high grit. Higher wealth was modestly associated with higher odds of high grit: highest tertile RRR = 1.17 (95% CI 1.00; 1.37), middle tertile not significant. Reporting a limiting illness was associated with lower odds of high grit (RRR = 0.78, 95% CI 0.68; 0.89). Better memory performance was modestly but significantly associated with high grit (RRR = 1.02, 95% CI 1.00; 1.04).

### Personality Grit

Table 5 shows the predictors of the grit-oriented personality component, explored simultaneously in the model.

**Table 5.**
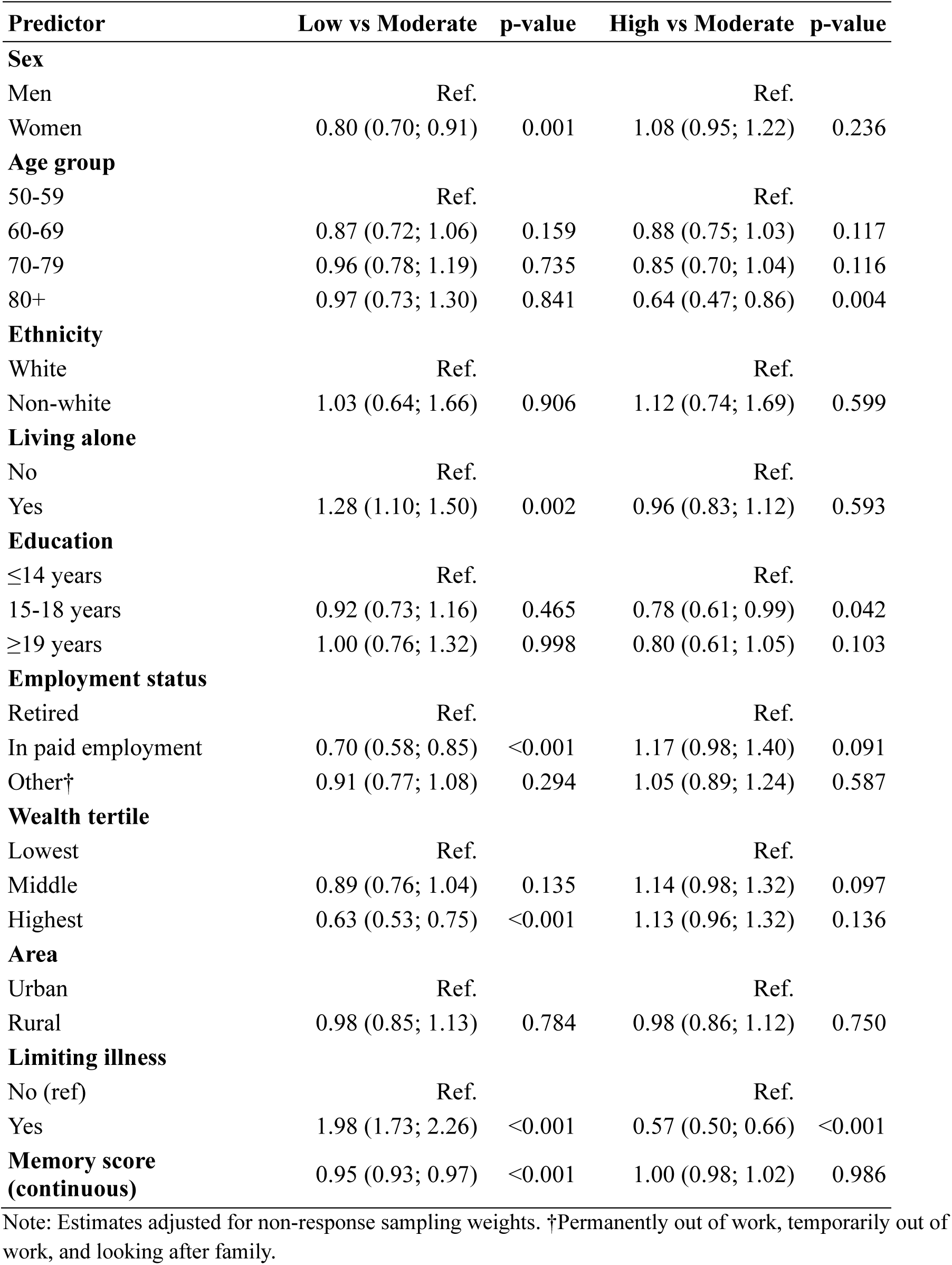
Predictors of the grit-oriented personality component, England 2008-2012.

#### Low personality grit versus moderate

After controlling for all other variables, women were significantly less likely than men to report low grit (RRR = 0.80, 95% CI 0.70; 0.91, p = 0.001). Living alone was associated with a higher likelihood of low grit (RRR = 1.28, 95% CI 1.10; 1.50). Being in paid employment was linked to a reduced risk of low grit relative to moderate grit (RRR = 0.70, 95% CI 0.58; 0.85), while higher wealth showed lower associations. Participants reporting a limiting illness were substantially more likely to have low grit (RRR = 1.98, 95% CI 1.73; 2.26). Better memory performance was associated with a lower likelihood of low grit (RRR = 0.95, 95% CI 0.93; 0.97). Age, ethnicity, education and living in urban areas were not significantly associated with low grit compared to medium grit.

#### High personality grit versus moderate

In the comparison of high versus moderate grit, most of the sample characteristics did not show significant associations. Age showed limited associations, though those aged 80+ were significantly less likely to report high grit (RRR = 0.64, 95% CI 0.47; 0.86, p = 0.004). Participants with 15-18 years of education had slightly lower odds of high grit compared with those with fewer than 14 years (RRR = 0.78, 95% CI 0.61; 0.99, p = 0.042), while other education categories showed no associations. Reporting a limiting illness was strongly associated with lower odds of high grit (RRR = 0.57, 95% CI 0.50; 0.66).

Supplementary Tables 4-8 present comparisons of high versus low categories for each component. Across constructs, these analyses were broadly consistent with the main models: higher levels of growth mindset and grit were more likely among individuals with greater wealth, paid employment, and better memory performance, whereas older age (particularly 80+), living alone, and limiting longstanding illness were associated with a lower likelihood of belonging to the high group. These contrasts further emphasise the social and health patterning of lower psychological resources in later life.

## Discussion

Using nationally representative data from older adults in England, this study provides a population-level examination of Growth Mindset and Grit in later life. By operationalising proxy indicators across educational, personality, affective, and belief-based domains, we show that these constructs are not unitary traits but multidimensional psychological resources that are differentially distributed across socioeconomic position, health, and cognitive functioning. In doing so, we extend existing research beyond small or regionally specific samples and demonstrate that growth-oriented thinking and perseverance are socially patterned and embedded within cumulative life-course processes of advantage and disadvantage.

### Main findings

The distribution of the components revealed important differences between domains. The adult education component of Growth Mindset was heavily skewed towards the low category, with almost four fifths of participants reporting no engagement in formal learning over the previous four years. In contrast, the personality and belief based components of Growth Mindset showed more even distributions, with moderate and high scores well represented. A similar pattern emerged for the two Grit components, where personality based grit displayed a balanced distribution and affect based grit had an even slightly larger proportion of individuals in the high category. These results suggest that although formal educational activity becomes relatively uncommon in later adulthood, attitudinal, personality based and affective expressions of growth orientation and perseverance remain diverse in this population. This distinction underscores the importance of separating opportunity-dependent indicators from more dispositional or belief-based domains when studying psychological resources in older cohorts. It should be noted, however, that the categorisation of personality and belief components into Low, Moderate, and High groups is based on ±0.5 SD thresholds, which are somewhat arbitrary and do not represent empirically derived cut-points. As such, these categories are intended to aid interpretation rather than indicate qualitatively distinct subgroups.

Age gradients were particularly marked for the education and beliefs components of Growth Mindset. Older participants were substantially more likely to fall into the low categories of these domains and less likely to be classified as high. This was confirmed in the regression models, which showed that each older age group had increasingly higher risks of low adult education Growth Mindset and lower likelihoods of high belief based Growth Mindset. The personality component showed only modest age differences, which is consistent with the idea that trait based aspects of growth orientation may be more stable across the life course. This pattern aligns with life-course perspectives suggesting relative stability in core personality traits alongside structural shifts in opportunity structures with advancing age.

Age effects on Grit were more nuanced. Older age group were generally less likely to have low affect based Grit, suggesting that emotional steadiness, determination and positive affect related to perseverance may be relatively preserved. At the same time, the oldest participants were less likely to be in the high category of personality based Grit, which may reflect accumulated health difficulties or reductions in the energy required for sustained goal pursuit. Such findings are consistent with theories of selective optimisation with compensation, whereby goal persistence may be recalibrated in response to age-related constraints rather than simply diminished.

Gender differences were modest but consistent across some components. Women were less likely than men to fall into the low category of adult education Growth Mindset, but showed higher risks of low scores in the personality-based component. In contrast, women were less likely to have low personality based Grit and more likely to have high affect based Grit. These patterns may reflect gendered life-course experiences that shape how perseverance and growth-oriented beliefs are expressed, rather than inherent differences in psychological capacity. These findings are consistent with evidence that older women often display greater emotional resilience and social engagement, even when they have had fewer formal educational opportunities earlier in life (Phillips et al., 2016).

Living arrangements and health status emerged as central correlates for several components. Individuals living alone were more likely to fall into the low categories of the beliefs component of Growth Mindset and both Grit components, and were less likely to report high beliefs based Growth Mindset. Health status emerged as one of the most structurally consequential correlates across models. It was associated with substantially higher risks of low Growth Mindset and low Grit and markedly lower risks of high scores. These associations are consistent with studies showing that chronic illness and social isolation undermine perceived control, self-efficacy and motivation in later life and reduce opportunities for cognitively and emotionally stimulating activity (Bandura, 1997; Dickens et al., 2011; Kempen et al., 2006).

Socioeconomic position, captured through educational attainment, employment status, and household wealth, was consistently associated with more favourable profiles. Completing full-time education at age 19 years or older was associated with lower risks of low Growth Mindset for some components, particularly the personality-based domain, and with higher belief-based Growth Mindset. Being in paid employment, compared with being retired, was associated with lower odds of low adult education Growth Mindset, lower odds of low belief-based Growth Mindset, and reduced risks of low Grit in both affective and personality components. Household wealth showed particularly strong and consistent associations: higher wealth was linked to lower likelihoods of low Growth Mindset beliefs and low Grit, and to higher probabilities of belonging to the high categories of beliefs and Grit. Collectively, these findings suggest that Growth Mindset and Grit are socially patterned psychological resources, shaped by cumulative socioeconomic advantage across the life course. Rather than existing independently of structural conditions, these constructs appear embedded within material, educational, and occupational trajectories.

Cognitive performance also showed important associations with Growth Mindset and Grit. Higher memory scores were associated with more favourable profiles across most components, including lower odds of low Growth Mindset beliefs and low Grit, and higher likelihoods of high Growth Mindset beliefs and high Grit. Associations with the personality-based Growth Mindset component were more selective, but memory remained a consistent predictor of high scores across several domains. Although causal direction cannot be established, the observed associations raise the possibility of reciprocal relationships between cognitive functioning and growth-oriented psychological resources across the life course (Lachman et al., 2010). The strong associations observed between personality-based components and memory performance are consistent with evidence that personality traits related to flexibility, persistence, and emotional regulation are robust correlates of cognitive functioning in older adulthood (Luchetti et al., 2016).

Ethnicity and area-level context played a more limited role. After adjustment for socioeconomic and health factors, ethnicity was not independently associated with most components, suggesting that structural inequalities may operate primarily through material and health pathways rather than through ethnicity per se in this cohort.

### Comparisons with previous studies

Although direct comparisons with previous studies are limited by differences in measurement and study design, the present findings are in line with evidence suggesting that growth-oriented beliefs and behaviours tend to co-occur with more favourable cognitive, health, and social profiles in later life (Fabio et al., 2023; Lachman & Agrigoroaei, 2010). Our findings extend prior work by demonstrating that these associations are observable at the population level and persist even when constructs are operationalised using indirect indicators. However, the steep age gradients and high concentration of low scores observed particularly for the adult education component highlight the limitations of applying education based Growth Mindset proxies to older populations. This likely reflects structural barriers to formal learning in later adulthood, rather than a lack of belief in personal growth. Studies of adult learning show that older adults often participate in informal learning rather than formal courses (Schuller et al., 2009), suggesting that measures of Growth Mindset in later life should be broadened to capture self-directed learning, technological adaptation and everyday problem solving.

Our findings are also broadly consistent with studies showing associations between grit and more favourable cognitive, health and social outcomes (Fleck et al., 2024; Kim & Lee, 2015; Rhodes & Giovannetti, 2022). The present analyses extend this literature by showing that different components of grit are shaped by different socio demographic factors. For example, Rhodes and Giovannetti reported no association between grit and age, yet we found that the oldest participants were less likely to reach the highest levels of personality based grit, although they were not over represented in the low category. This divergence may reflect differences in sample composition or cultural context and could indicate that the highest levels of sustained effort become harder to maintain in late life when health burdens accumulate.

A central contribution of this study is the demonstration that treating Growth Mindset and Grit as multidimensional constructs reveals heterogeneity that would remain obscured under single-trait conceptualisations. Much prior research has conceptualised Growth Mindset primarily as beliefs about the malleability of intelligence, as originally articulated in theories of implicit intelligence (Burnette et al., 2013; Dweck, 2006). These findings challenge intelligence-centred conceptualisations of Growth Mindset and suggest that growth orientation in later life may be better understood through belief systems, emotional regulation, and adaptive persistence in the face of structural and health-related constraints.

Personality based and belief based components displayed richer variation and stronger associations with social and health characteristics than the education based component, which was heavily influenced by opportunity structures. Similarly, distinguishing between affective and personality based grit highlighted important differences in the socio demographic and health correlates of perseverance. These findings indicate that growth orientation and grit in later life may be expressed less through formal learning or intelligence and more through everyday attitudes, emotional regulation and the capacity to persist in the face of health and social challenges.

### Strengths and limitations

A strength of this study is the use of a large, nationally representative cohort of older adults, which provides robust population level estimates of how different aspects of Growth Mindset and Grit are distributed in later life. A possible limitation is that the measures used were proxy indicators rather than validated scales, and although they captured meaningful variation, they do not fully reflect the theoretical scope of Growth Mindset or Grit. Furthermore, the study used cross-sectional data which do not allow for examination of changes in Growth Mindset or Grit over time.

In conclusion, this study demonstrates that Growth Mindset and Grit in later life are multidimensional and socially structured psychological resources, closely intertwined with socioeconomic position, health, and cognitive functioning. These findings move the field beyond small-scale cross-sectional studies and situate growth-oriented beliefs and perseverance within broader life-course and inequality frameworks. As populations age, understanding how such psychological resources are shaped by structural conditions may be critical for designing interventions that promote equitable trajectories of healthy ageing.

## Data Availability

The data that support the findings of this study are available from the UK Data Service (English Longitudinal Study of Ageing), subject to registration and standard access conditions.

## Supplementary Materials

Below, we describe the operationalisation of each component of growth mindset and grit. The items included in each component are described below and in STable 1; they were selected using a theory-informed, conceptually driven approach, guided by prior research on growth mindset, perseverance, and traits associated with openness to change and intellectual engagement. Selection was based on conceptual relevance and face validity, ensuring that each item reflected the underlying psychological construct of interest.

### Growth Mindset Personality

Personality was assessed at Wave 5 (2010-2011) using a short form of the Midlife Development Inventory (MIDI) (Lachman & Weaver, 1997), an instrument also employed in the US Health and Retirement Study. The questionnaire asked participants to rate how well a series of 26 descriptive adjectives described them, using a four-point Likert scale (1 = a lot, 4 =not at all). To capture trait-based tendencies consistent with openness to change and intellectual engagement, we constructed a personality index based on the following five items: Curious, Imaginative, Intelligent, Creative, and Broad-minded. To ensure that higher scores reflected greater endorsement of the trait, all items were reverse-coded. Each reverse-coded item was then standardised (z-scored) to place them on a common metric (mean = 0, SD = 1). A continuous growth-oriented personality score was constructed by averaging the five standardised items, such that higher values represented stronger endorsement of growth-related traits (Mean −0.000493 S.D. 0.69825 range −3.2876 to 1.3818) To aid interpretation, this continuous score was then categorised into three groups based on ±0.5 standard deviation thresholds: Low (< −0.5 SD), Moderate (−0.5 to 0.5 SD), and High (> 0.5 SD).

### Growth Mindset Beliefs

Growth mindset beliefs were operationalised using three items (“Chooses to do things they have never done”, “Feels that life is full of opportunities”, “Feels the future looks good to them”) from the CASP-19 (Hyde et al., 2003) quality of life questionnaire, administered at each ELSA wave, to capture attitudes consistent with growth-oriented perspectives. CASP19 has been administered at every wave via a self-completion questionnaire, with responses using a four-point Likert scale (1 = often, 4 =never). Responses were first recoded so that higher values reflected stronger endorsement of growth mindset beliefs. Each item was then standardised (z-scored) to place them on a common metric (mean = 0, SD = 1). A composite proxy index was created by averaging the three standardised items, yielding a continuous score of growth mindset belief (Mean −0.00128 S.D. 0.82654 range −2.4842 to 1.76625), with higher values representing stronger endorsement. The continuous score was categorised into three groups using ±0.5 standard deviation thresholds: Low (< −0.5 SD), Moderate (−0.5 to 0.5 SD), and High (> 0.5 SD).

### Growth Mindset Adult Learning

Engagement in formal adult learning activities across waves was used as a behavioural indicator of growth-oriented learning within institutional contexts. At each wave, respondents were asked to report on their participation in formal learning experiences. Specifically, three items were used to capture continued learning and personal development: (1) whether they had obtained a further qualification in the past two years, (2) whether they had attended a formal course in the past month, and (3) whether they had participated in formal education or training in the past twelve months. These indicators reflect observable engagement in structured learning environments, which may signal an orientation toward continued learning and development. For this component, we extended the time window to include both Waves 4 and 5 of ELSA (2008-09 and 2010-12, respectively) rather than relying solely on Wave 5. This approach increases the likelihood of capturing infrequent or episodic learning behaviours that may not occur in a single wave, providing a more comprehensive measure of participants’ engagement in growth-oriented educational activity. It also reduces the risk of underestimating learning engagement due to the timing of data collection, ensuring that the resulting score better reflects participation in formal learning over time. A growth-oriented learning categorical variable was computed from the information of these variables as follows: Low (if they did not engage in any of the three learning activities in the 4 years prior the interview), Moderate (if they engaged in either a formal course or formal education training at least once in the 4 years prior the interview), High (if they obtained a further qualification in the 4 years prior the interview).

### Grit Personality

Personality-based grit was operationalised using selected items from the Midlife Development Inventory (MIDI) personality questionnaire administered at Wave 5 (2010-11). This approach captures grit indirectly through stable personality characteristics, rather than using a dedicated grit scale.

The rationale for the selected traits is grounded in the core components of grit: perseverance, resilience, focus, and diligence. Specifically, the following MIDI items were chosen to reflect these dimensions: *Hardworking* and *Responsible* (perseverance of effort), *Calm* (resilience), *Organised* (focus), *Active* (commitment to goals), and *Thorough* (diligence/practice). These traits collectively capture an individual’s tendency to maintain effort and interest over long-term goals. Participants were asked to record how well each of the item described them, responses were recorded on a 5-point Likert scale ranging from 1 = “a lot” to 4 = “not at all.

Each item was reverse-coded where necessary so that higher values reflected stronger endorsement of grit-related characteristics. Items were then standardised (z-scored) to place them on a common metric (mean = 0, SD = 1), and a composite personality-based grit score was calculated by averaging the standardised items (Mean −0.000434 S.D. 0.675 range −2.952 to 1.196). Higher values indicate greater expression of grit. The continuous score was categorised into three groups based on ±0.5 standard deviation thresholds: Low (< −0.5 SD), Moderate (−0.5 to 0.5 SD), and High (> 0.5 SD).

### Grit Affect

Affect-based grit was operationalised using selected positive affect items from a self-completion questionnaire administered in Wave 5 (2010-11). The questionnaire used measures of positive and negative affect chosen from Positive and Negative Affect Schedule Expanded Form (Watson & Clark, 1994) (PANAS-X; Watson & Clark, 1994) also administered in the US Health and Retirement Study. The items assess the frequency and intensity of both positive emotions (e.g., enthusiasm, alertness) and negative emotions (e.g., distress, irritability), providing a reliable overview of participants’ recent emotional experiences. Participants were asked to record to what degree in the past 30 days, they experienced 13 feelings, responses were recorded on a 5-point Likert scale ranging from 1 = “very much” to 5 = “not at all. Specifically, the following items were chosen to reflect these dimensions: *Determined*, *Hopeful*, *Attentive*, *Interested*, and *Inspired*. The rationale for the selected affects is grounded in the motivational and engagement aspects of grit. These positive affects collectively capture an individual’s tendency to maintain focus, energy, and optimism in the pursuit of long-term goals.

Each item was reverse-coded so that higher values reflected greater positive affect, indicative of stronger grit-related tendencies. Items were then standardised (z-scored) to place them on a common metric (mean = 0, SD = 1), and a composite affect-based grit score was calculated by averaging the standardised items (Mean −0.00210 S.D. 0.83 range −3.314 to 1.581). Higher values indicate greater expression of grit through positive affect. The continuous score was categorised into three groups based on ±0.5 standard deviation thresholds: Low (< −0.5 SD), Moderate (−0.5 to 0.5 SD), and High (> 0.5 SD).

---

**Table S1.** Components of each Growth Mindset and Grit Measure.

| Construct | Source / Questionnaire | Wave(s) | Components / Items | Description |
| --- | --- | --- | --- | --- |
| Growth Mindset Adult Learning | Main questionnaire items | Waves 4 (2008-09)<br>Wave 5 (2010-12) | - Obtained a further qualification in the past two years- Attended a formal course in the past month- Participated in formal education or training in the past 12 months | Behavioural indicator of growth-oriented learning; captures participation in formal learning and personal development, reflecting an individual's orientation toward continued learning and self-improvement |
| Growth Mindset Personality | Self-completion questionnaire using Midlife Development Inventory (MIDI) | Wave 5 (2010-11) | - Curious- Imaginative- Intelligent- Creative- Broad-minded | Assesses trait-based tendencies consistent with openness to change and intellectual engagement; participants rated 26 adjectives on a 4-point Likert scale (1 = a lot, 4 = not at all) |
| Growth Mindset Beliefs | Self-completion questionnaire using CASP-19 quality of life questionnaire | Wave 5 (2012-13) | - “Chooses to do things they have never done”- “Feels that life is full of opportunities”- “Feels the future looks good to them” | Operationalises growth mindset attitudes; CASP-19 administered via self-completion questionnaire at every wave, responses on a 4-point Likert scale (1 = often, 4 = never) |
| Grit Personality | Self-completion questionnaire using Midlife Development Inventory (MIDI) | Wave 5 (2010-11) | - Hardworking and Responsible (perseverance of effort)- Calm (resilience)- Organised (focus)- Active (commitment to goals)- Thorough (diligence/practice) | Personality-based indicator of grit; selected items capture perseverance, resilience, focus, and diligence; participants rated how well each item described them on a 5-point Likert scale (1 = “a lot”, 4 = “not at all”) |
| Grit Positive Affect | Self-completion questionnaire using Positive and Negative Affect Schedule Expanded Form (PANAS-X) | Wave 5 (2010-11) | - Determined- Hopeful- Attentive- Interested- Inspired | Affect-based indicator of grit; captures motivation, interest, and sustained engagement; participants reported frequency/intensity of emotions over past 30 days on a 5-point Likert scale (1 = “very much”, 5 = “not at all”) |

**Table S2.** Distribution of the components of the Growth Mindset and Grit scores, England 2008-2012.

|  | **Low**<br>**% (95%CI)** | **Moderate**<br>**% (95%CI)** | **High**<br>**% (95%CI)** |
| --- | --- | --- | --- |
| **Growth Mindset** |  |  |  |
| **Education** | 79.3 | 12.7 | 8.1 |
|  | (78.3; 80.2) | (11.9; 13.5) | (7.4; 8.8) |
| *N* | 5971 | 976 | 566 |
| **Personality** | 26.3 | 49.9 | 23.8 |
|  | (25.2; 27.4) | (48.7; 51.1) | (22.8; 24.8) |
| *N* | 1832 | 3684 | 1817 |
| **Beliefs** | 27.7 | 37.4 | 34.9 |
|  | (26.6; 28.9) | (36.2; 38.6) | (33.7; 36.1) |
| *N* | 1893 | 2763 | 2677 |
| **Grit** |  |  |  |
| **Personality** | 24.1 | 44.7 | 28.2 |
|  | (23.1; 25.2) | (46.5; 48.9) | (27.1; 29.3) |
| *N* | 1669 | 3558 | 2106 |
| **Affect** | 26.7 | 44.5 | 28.8 |
|  | (25.6; 27.9) | (43.3; 45.7) | (27.7; 29.9) |
| *N* | 1854 | 3268 | 2211 |

Note: Estimates adjusted for non-response sampling weights

**Table S3.** Distribution of the components of the Growth Mindset and Grit scores according to age, England 2008-2012.

|  | **Low**<br>**Mean (s.e.)** | **Moderate**<br>**Mean (s.e.)** | **High**<br>**Mean (s.e.)** |
| --- | --- | --- | --- |
| **Growth Mindset Education** | 68.6 (0.15) | 62.4 (0.26) | 60.3 (0.26) |
| **Growth Mindset Personality** | 68.6 (0.27) | 67.0 (0.19) | 66.0 (0.26) |
| **Growth Mindset Beliefs** | 70.0 (0.29) | 66.9 (0.21) | 65.1 (0.18) |
| **Grit Personality** | 68.1 (0.29) | 67.1 (0.20) | 66.3 (0.22) |
| **Grit Affect** | 69.5 (0.29) | 67.0 (0.19) | 65.5 (0.24) |

Note: Estimates adjusted for non-response sampling weights. Sample size unweighted.

**Table S4.** Predictors of the growth mindset-oriented adult education component, England 2008-2012.

| **Predictor** | **High vs Low** | **p-value** |
| --- | --- | --- |
| **Sex** |  |  |
| Men | Ref. |  |
| Women | 1.03 (0.85; 1.26) | 0.760 |
| **Age group** |  |  |
| 50-59 | Ref. |  |
| 60-69 | 0.56 (0.45; 0.69) | <0.001 |
| 70- 79 | 0.17 (0.12; 0.26) | <0.001 |
| 80+ | 0.11 (0.05; 0.23) | <0.001 |
| **Ethnicity** |  |  |
| White | Ref. |  |
| Non-white | 1.03 (0.54; 1.95) | 0.933 |
| **Living alone** |  |  |
| No | Ref. |  |
| Yes | 1.04 (0.80; 1.36) | 0.754 |
| **Education** |  |  |
| ≤14 years | Ref. |  |
| 15-18 years | 0.84 (0.50; 1.40) | 0.499 |
| ≥19 years | 1.24 (0.72; 2.13) | 0.442 |
| **Employment status** |  |  |
| Retired | Ref. |  |
| In paid employment | 4.43 (2.96; 6.62) | <0.001 |
| Other† | 2.01 (1.34; 3.02) | 0.001 |
| **Wealth tertile** |  |  |
| Lowest | Ref. |  |
| Middle | 1.08 (0.85; 1.38) | 0.520 |
| Highest | 0.99 (0.76; 1.29) | 0.931 |
| **Area** |  |  |
| Urban | Ref. |  |
| Rural | 0.94 (0.76; 1.16) | 0.559 |
| **Limiting illness** |  |  |
| No | Ref. |  |
| Yes | 0.84 (0.66; 1.06) | 0.140 |
| **Memory score (continuous)** | 1.04 (1.01; 1.08) | 0.006 |

Note: Estimates adjusted for non-response sampling weights. †Permanently out of work, temporarily out of work, and looking after family.

**Table S5.** Predictors of the growth mindset-oriented personality component, England 2008-2012.

| **Predictor** | **High vs Low** | **p-value** |
| --- | --- | --- |
| **Sex** |  |  |
| Men | Ref. |  |
| Women | 0.80 (0.69; 0.93) | 0.004 |
| **Age group** |  |  |
| 50-59 | Ref. |  |
| 60-69 | 1.05 (0.86; 1.29) | 0.612 |
| 70-79 | 0.99 (0.78; 1.26) | 0.945 |
| 80+ | 0.92 (0.66; 1.30) | 0.646 |
| **Ethnicity** |  |  |
| White | Ref. |  |
| Non-white | 0.81 (0.48; 1.34) | 0.406 |
| **Living alone** |  |  |
| No | Ref. |  |
| Yes | 1.00 (0.84; 1.19) | 0.964 |
| **Education** |  |  |
| ≤14 years | Ref. |  |
| 15-18 years | 1.06 (0.81; 1.39) | 0.677 |
| ≥19 years | 1.96 (1.43; 2.68) | <0.001 |
| **Employment status** |  |  |
| Retired | Ref. |  |
| In paid employment | 1.24 (1.00; 1.55) | 0.050 |
| Other† | 1.06 (0.87; 1.28) | 0.575 |
| **Wealth tertile** |  |  |
| Lowest | Ref. |  |
| Middle | 1.06 (0.89; 1.27) | 0.496 |
| Highest | 1.20 (0.99; 1.45) | 0.058 |
| **Area** |  |  |
| Urban | Ref. |  |
| Rural | 1.14 (0.97; 1.34) | 0.124 |
| **Limiting illness** |  |  |
| No | Ref. |  |
| Yes | 0.77 (0.66; 0.91) | 0.001 |
| **Memory score (continuous)** | 1.10 (1.07; 1.12) | <0.001 |

Note: Estimates adjusted for non-response sampling weights. †Permanently out of work, temporarily out of work, and looking after family.

**Table S6.** Predictors of the growth mindset-oriented beliefs component, England 2008-2012.

| **Predictor** | **High vs Low** | **p-value** |
| --- | --- | --- |
| **Sex** |  |  |
| Men | Ref. |  |
| Women | 1.18 (1.02; 1.36) | 0.023 |
| **Age group** |  |  |
| 50-59 | Ref. |  |
| 60-69 | 1.30 (1.07; 1.58) | 0.007 |
| 70-79 | 0.97 (0.77; 1.21) | 0.768 |
| 80+ | 0.58 (0.42; 0.81) | 0.001 |
| **Ethnicity** |  |  |
| White | Ref. |  |
| Non-white | 1.20 (0.69; 2.10) | 0.521 |
| **Living alone** |  |  |
| No | Ref. |  |
| Yes | 0.62 (0.53; 0.73) | <0.001 |
| **Education** |  |  |
| ≤14 years | Ref. |  |
| 15-18 years | 1.01 (0.77; 1.32) | 0.952 |
| ≥19 years | 1.34 (0.98; 1.84) | 0.069 |
| **Employment status** |  |  |
| Retired | Ref. |  |
| In paid employment | 1.34 (1.09; 1.66) | 0.005 |
| Other† | 0.99 (0.82; 1.19) | 0.903 |
| **Wealth tertile** |  |  |
| Lowest | Ref. |  |
| Middle | 1.46 (1.24; 1.73) | <0.001 |
| Highest | 2.42 (2.02; 2.91) | <0.001 |
| **Area** |  |  |
| Urban | Ref. |  |
| Rural | 1.11 (0.95; 1.29) | 0.193 |
| **Limiting illness** |  |  |
| No | Ref. |  |
| Yes | 0.28 (0.24; 0.32) | <0.001 |
| **Memory score (continuous)** | 1.07 (1.05; 1.09) | <0.001 |

Note: Estimates adjusted for non-response sampling weights. †Permanently out of work, temporarily out of work, and looking after family.

**Table S7.**
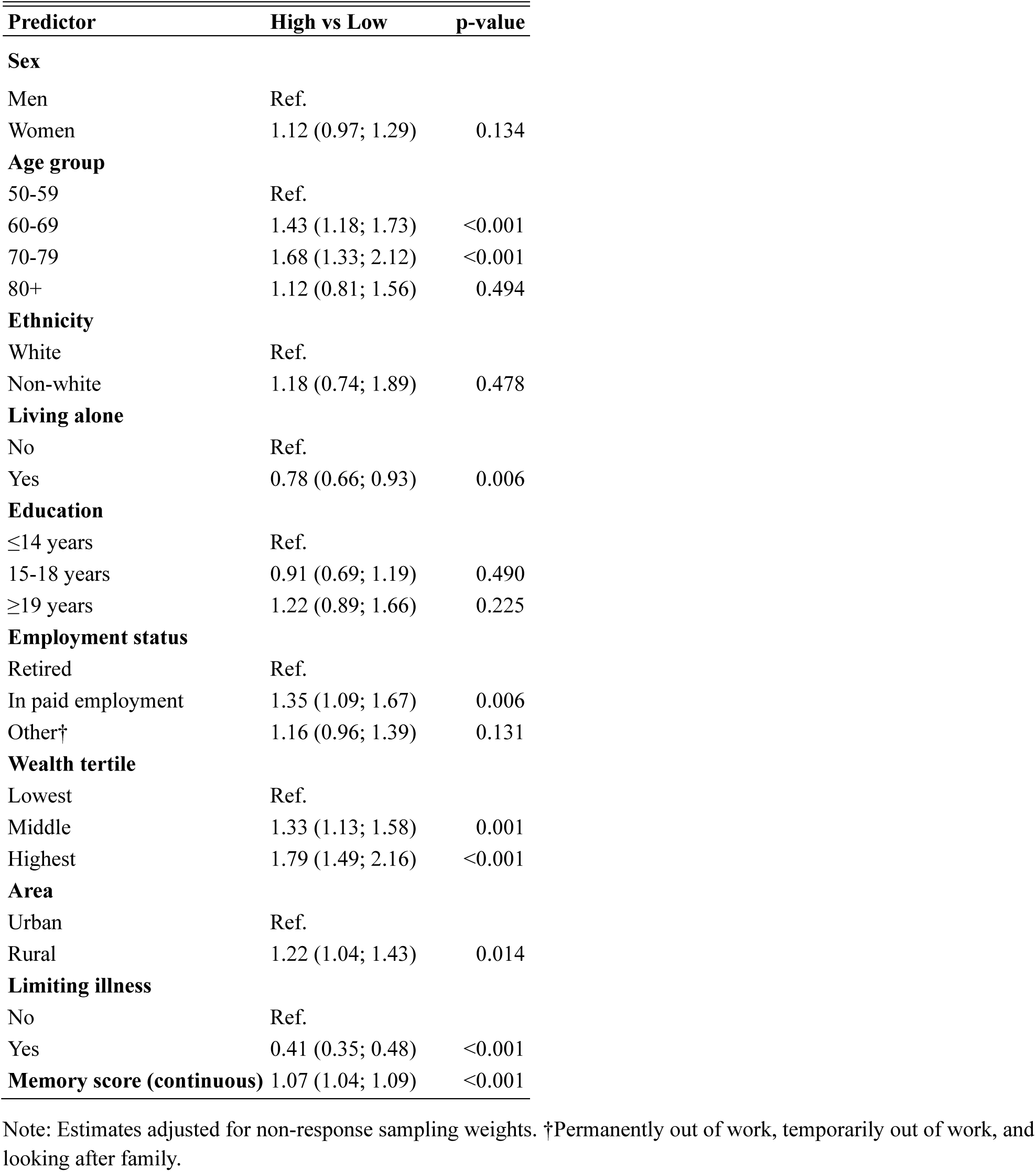
Predictors of the grit-oriented affect component, England 2008-2012.

**Table S8.** Predictors of the grit-oriented personality component, England 2008-2012.

| **Predictor** | **High vs Low** | **p-value** |
| --- | --- | --- |
| **Sex** |  |  |
| Men | Ref. |  |
| Women | 1.35 (1.16; 1.57) | <0.001 |
| **Age group** |  |  |
| 50-59 | Ref. |  |
| 60-69 | 1.01 (0.82; 1.24) | 0.912 |
| 70-79 | 0.89 (0.70; 1.13) | 0.326 |
| 80+ | 0.66 (0.47; 0.94) | 0.020 |
| **Ethnicity** |  |  |
| White | Ref. |  |
| Non-white | 1.09 (0.66; 1.80) | 0.748 |
| **Living alone** |  |  |
| No | Ref. |  |
| Yes | 0.75 (0.63; 0.89) | 0.001 |
| **Education** |  |  |
| ≤14 years | Ref. |  |
| 15-18 years | 0.85 (0.65; 1.12) | 0.245 |
| ≥19 years | 0.80 (0.58; 1.10) | 0.168 |
| **Employment status** |  |  |
| Retired | Ref. |  |
| In paid employment | 1.66 (1.33; 2.07) | <0.001 |
| Other† | 1.14 (0.94; 1.39) | 0.172 |
| **Wealth tertile** |  |  |
| Lowest | Ref. |  |
| Middle | 1.28 (1.07; 1.53) | 0.007 |
| Highest | 1.78 (1.46; 2.17) | <0.001 |
| **Area** |  |  |
| Urban | Ref. |  |
| Rural | 1.00 (0.85; 1.18) | 0.994 |
| **Limiting illness** |  |  |
| No | Ref. |  |
| Yes | 0.29 (0.25; 0.34) | <0.001 |
| **Memory score (continuous)** | 1.05 (1.03; 1.08) | <0.001 |

Note: Estimates adjusted for non-response sampling weights. †Permanently out of work, temporarily out of work, and looking after family.

## References

Bandura, A. (1997). Self-efficacy : the exercise of control. W.H. Freeman.

Burnette, J. L., O’Boyle, E. H., VanEpps, E. M., Pollack, J. M., & Finkel, E. J. (2013, May). Mind-sets matter: a meta-analytic review of implicit theories and self-regulation. Psychol Bull, 139(3), 655–701. 10.1037/a0029531

Dickens, A. P., Richards, S. H., Greaves, C. J., & Campbell, J. L. (2011, Aug 15). Interventions targeting social isolation in older people: a systematic review. BMC Public Health, 11, 647. 10.1186/1471-2458-11-647

Duckworth, A. (2016). Grit : the power of passion and perseverance (First Scribner hardcover edition. ed.). Scribner.

Dweck, C. S. (2006). Mindset : the new psychology of success (1st ed.). Random House. Contributor biographical information http://www.loc.gov/catdir/enhancements/fy0624/2005046454-b.html

Publisher description http://www.loc.gov/catdir/enhancements/fy0624/2005046454-d.html Sample text http://www.loc.gov/catdir/enhancements/fy0624/2005046454-s.html

Dweck, C. S., & Yeager, D. S. (2019, May). Mindsets: A View From Two Eras. Perspect Psychol Sci, 14(3), 481–496. 10.1177/1745691618804166

Fabio, R. A., Chiatto, L. M., & Colombo, B. (2023). Growth mind-set and healthy behaviors in older adults. Activities, Adaptation & Ageing, 48(4), 613–626. 10.1080/01924788.2023.2252249

Fleck, J. I., Patel, P., Riley, E., & Ferri, C. V. (2024, May). Mindset matters: contributions from grit and growth mindsets to successful aging. Aging Ment Health, 28(5), 819–827. 10.1080/13607863.2023.2280626

Heslin, P. A., L., B. J., & Ryu, N. G. (2021). Does a Growth Mindset Enable Successful Aging? Work, Aging and Retirement, 7(2), 11. 10.1093/workar/waaa029

Hyde, M., Wiggins, R. D., Higgs, P., & Blane, D. B. (2003, May). A measure of quality of life in early old age: the theory, development and properties of a needs satisfaction model (CASP-19). Aging Ment Health, 7(3), 186–194. 10.1080/1360786031000101157

Kempen, G. I., Ranchor, A. V., van Sonderen, E., van Jaarsveld, C. H., & Sanderman, R. (2006, Mar). Risk and protective factors of different functional trajectories in older persons: are these the same? J Gerontol B Psychol Sci Soc Sci, 61(2), P95–101. 10.1093/geronb/61.2.p95

Kim, Y. J., & Lee, C. S. (2015). Effects of grit on the successful ageing of the elderly in Korea. Indian Journal of Science and Technology, 8(S7), 373–378. 10.17485/ijst/2015/v8iS7/70421

Lachman, M. E., & Agrigoroaei, S. (2010, Oct 11). Promoting functional health in midlife and old age: long-term protective effects of control beliefs, social support, and physical exercise. PLoS One, 5(10), e13297. 10.1371/journal.pone.0013297

Lachman, M. E., & Weaver, S. L. (1997). The Midlife Development Inventory (MIDI) Personality Scales: Scale construction and scoring. Brandeis University. https://www.brandeis.edu/psychology/lachman/pdfs/midi-personality-scales.pdf

Leanos, S., Kurum, E., Strickland-Hughes, C. M., Ditta, A. S., Nguyen, G., Felix, M., Yum, H., Rebok, G. W., & Wu, R. (2023, Aug 2). The Impact of Learning Multiple Real-World Skills on Cognitive Abilities and Functional Independence in Healthy Older Adults. J Gerontol B Psychol Sci Soc Sci, 78(8), 1305–1317. 10.1093/geronb/gbad053

Luchetti, M., Terracciano, A., Stephan, Y., & Sutin, A. R. (2016, Jul). Personality and Cognitive Decline in Older Adults: Data From a Longitudinal Sample and Meta-Analysis. J Gerontol B Psychol Sci Soc Sci, 71(4), 591–601. 10.1093/geronb/gbu184

Rhodes, E., & Giovannetti, T. (2022, Jun). Grit and successful aging in older adults. Aging Ment Health, 26(6), 1253–1260. 10.1080/13607863.2021.1919990

Schuller, T., Watson, D., & National Institute of Adult Continuing Education (England and Wales). (2009). Learning through life : inquiry into the future for lifelong learning. National Institute of Adult Continuing Education.

Sheffler, P., Kurum, E., Sheen, A. M., Ditta, A. S., Ferguson, L., Bravo, D., Rebok, G. W., Strickland-Hughes, C. M., & Wu, R. (2023, Jun). Growth Mindset Predicts Cognitive Gains in an Older Adult Multi-Skill Learning Intervention. Int J Aging Hum Dev, 96(4), 501–526. 10.1177/00914150221106095

Steptoe, A., Breeze, E., Banks, J., & Nazroo, J. (2013, Dec). Cohort profile: the English longitudinal study of ageing. Int J Epidemiol, 42(6), 1640–1648. 10.1093/ije/dys168

Watson, D., & Clark, L. A. (1994). The PANAS-X: Manual for the Positive and Negative Affect Schedule - Expanded Form. https://iro.uiowa.edu/esploro/outputs/other/The-PANAS-X-Manual-for-the-Positive/9983557488402771

